# Government health care worker training needs for schistosomiasis morbidity management

**DOI:** 10.64898/2026.03.13.26348332

**Authors:** Phyllis Munyiva Isaiah, Betty Nabatte, Lauren Wilburn, John Bosco Oryema, Noah Ukumu, Morris Okumu, Juma Nabhonge, Hilda Kyarisiima, Prudence Beinamaryo, Victor Anguajibi, Christopher K. Opio, Narcis B. Kabatereine, Goylette F. Chami

**Affiliations:** Big Data Institute, Nuffield Department of Population Health, University of Oxford, Oxford, United Kingdom; Division of Vector-Borne and Neglected Tropical Diseases Control, Uganda Ministry of Health, Kampala, 15 Bombo Road, Kampala, Uganda; Pakwach District Local Government, Uganda Ministry of Health, Pakwach Town, Uganda; Buliisa District Local Government, Uganda Ministry of Health, Buliisa Town, Uganda; Mayuge District Local Government, Uganda Ministry of Health, Mayuge Town, Uganda; Uganda Institute of Allied Health Sciences, Kampala, Uganda; Aga Khan University Hospital, 3rd Parkland Avenue, PO Box 30270-00100, Nairobi, Kenya

**Keywords:** Schistosomiasis, morbidity, disability, mortality, health system, capacity, strengthening, training, case management, hepatosplenic, Africa

## Abstract

**Background:** Schistosomiasis causes substantial chronic morbidity in sub-Saharan Africa, yet case definitions, clinical management guidance, and health worker training for schistosomiasis-related morbidity remain limited.

**Methodology:** We conducted a qualitative needs assessment for schistosomiasis morbidity management. Workshops were held over one day in each of Pakwach, Buliisa, and Mayuge Districts in Uganda in October 2024. 105 government health workers participated including clinicians, nurses, laboratory technicians, sonographers, and district health managers from health facilities at different levels of care. The workshops comprised six structured sessions: presentations on schistosomiasis burden in Uganda and the SchistoTrack cohort, a clinical case report by an expert clinician, an interactive session on patient case studies from the SchistoTrack cohort, mapping of patient pathways, anonymous participation and feedback, and demonstrations of schistosomiasis diagnosis. Workshop discussions were documented through notes taken in English and analysed using qualitative thematic analysis as per Braun and Clarke.

**Findings:** Health workers demonstrated substantial gaps in understanding schistosomiasis case definitions, particularly in distinguishing current infection from chronic morbidity and in grading disease severity. Patient pathways for schistosomiasis morbidity management were fragmented and inconsistent, with weak triage, unclear referral and feedback mechanisms, and limited follow-up across facility levels. Health facilities lacked essential capacity and resources, including routine access to praziquantel outside mass drug administration, diagnostic reagents, functional ultrasound equipment, trained sonographers, and standardized training and reference tools. Collectively, these gaps contributed to inconsistent clinical decision-making and under-recognition of severe schistosomiasis-related morbidity.

**Conclusions:** Integrating case management into routine health services through standardized case definitions, clearer patient pathways, and targeted practical training for health workers is essential to complement preventive chemotherapy and reduce preventable morbidity. The engagement framework and patient case studies used here can support needs-based assessments in other endemic settings to inform the development of context-appropriate clinical guidance and training programmes.

**Author summary:** Schistosomiasis is a disease common in sub-Saharan Africa that can cause serious, lifelong health problems, even after the infection itself has been treated. While mass drug administration programmes have reduced infections, we found that health facilities are often not equipped to recognize or manage the chronic illness that remains after mass drug administration. In this study, we worked with government health workers in three rural districts of Uganda to understand their experiences, challenges, and training needs related to schistosomiasis morbidity. Through interactive workshops, we learned that many health workers struggle to distinguish between current infection and long-term organ damage, especially liver disease. Patient care pathways were often unclear, referrals were poorly coordinated, and essential tools, such as diagnostic supplies, ultrasound access, and routine availability of treatment, were frequently missing. These gaps make it difficult to identify severe cases early and provide consistent care. We show that strengthening routine health services through clearer case definitions, better training, and more coordinated patient pathways is essential to reduce preventable suffering from schistosomiasis. Our approach offers a practical way to support health systems in other endemic settings facing similar challenges.

## Introduction

Schistosomiasis is a leading cause of disability and likely mortality in sub-Saharan Africa (SSA), yet there are no clinical management guidelines within endemic countries or from the World Health Organization (WHO). The two main species that infect humans in SSA, where nearly 90% of global cases occur, are *Schistosoma haematobium*, which causes urogenital schistosomiasis and *S. mansoni*, which causes intestinal schistosomiasis [1]. Schistosomiasis is estimated to cause 1.9 million disability-adjusted life years [2] and about 200,000 deaths annually [3], but this burden is likely underestimated due to limited disability and mortality data from SSA [4] and unaccounted for or undifferentiated morbidity attributable to schistosomiasis [5–7]. Of particular concern is morbidity due to *S. mansoni*, which has been shown to be less likely than morbidity due to *S. haematobium* to resolve after treatment [8].

Repeated infections with *S. mansoni* lead to hepatosplenic disease, characterized by the development of hepatic granulomas, which may first manifest clinically as enlargement of the liver and spleen with general symptoms of abdominal pain, possible anaemia, and other non-specific indications [9]. As the disease progresses, fibrosis is observed within the periportal regions via ultrasound [10]. Recent evidence suggests that periportal fibrosis is not amenable to treatment with praziquantel that is administered routinely through mass drug administration (MDA) for current infections [11]. The presentation of periportal fibrosis is complex. The presence of two or more chronic conditions (multimorbidity) is common with more than 50% of individuals in *S. mansoni*-endemic communities estimated to have hepatosplenic multimorbidity [12]. Individuals with periportal fibrosis have been shown to have concurrent conditions of hypersplenism, anaemia, ascites, gastro-oesophageal varices, splenic varices, portal vein enlargement, and gall bladder wall thickening [13, 14]. Despite the substantial disease burden that remains after MDA and its complex presentations, MDA remains as the sole strategy for schistosomiasis control.

MDA campaigns in most endemic countries are primarily supported by external donors and are implemented largely outside the routine health system [15]. Typically conducted once a year, these campaigns focus on school-going children, who are mainly reached through school-based delivery platforms. The strategy does not rely on government health workers; instead, primary school teachers and community (village) medicine distributors—both volunteer-based—implement MDA. Consequently, praziquantel is not supplied through health facilities, as donated praziquantel is allocated exclusively for vertical MDA programmes [16]. Hence, schistosomiasis surveillance, monitoring, and follow-up activities are not embedded within routine health services.

The goal of MDA is to control schistosomiasis by reducing infection intensities before disease develops [17]. As a result, clinical management guidelines for schistosomiasis within health facilities remain limited and there exist substantial gaps in health worker knowledge about schistosomiasis-related morbidities. Diagnosis and management is further complicated by the presence of other causes of liver disease such as alcohol use or hepatitis B that can be difficult to distinguish from schistosomiasis-related hepatosplenic disease [18]. The WHO schistosomiasis guidelines [1] and Neglected Tropical Disease Roadmap [17] currently focus on elimination of human schistosomiasis as a public health problem by 2030. Though these guidelines still focus on the reduction of current infection intensity, which has been repeatedly shown to be uncorrelated with current morbidity [11, 12], there have been recent calls by African experts for direct morbidity surveillance to eliminate severe schistosomiasis morbidity [19] and recent suggestions by the WHO of indicators [20] that might be recorded within routine health facilities. Yet, there are no clear case definitions for what constitutes severe schistosomiasis-related morbidity or how to train health workers to recognize and manage such cases.

We conducted a qualitative needs assessment by hosting workshops with a wide range of specialities of government health workers in schistosomiasis-endemic areas of Pakwach, Buliisa, and Mayuge Districts of Uganda. By raising awareness of severe schistosomiasis morbidity and identifying associated health system needs, the aim of the workshops was to identify feasible case management options within existing resource constraints and to provide information that can inform the development of a schistosomiasis-morbidity management training programme and future clinical guidelines.

## Methods

### Study context

This study was conducted within the catchment of the SchistoTrack Cohort, a prospective, community-based cohort in rural Uganda designed to improve understanding of schistosome transmission and morbidity in Pakwach, Buliisa, and Mayuge districts. Additional cohort details can be found elsewhere [21]. *S. mansoni* is highly prevalent in Uganda [22], with some SchistoTrack cohort villages reporting infection rates above 50% [12]. In 2022 at the cohort baseline, 12.1% of participants were diagnosed with periportal fibrosis, with the highest burden in Pakwach (19.5%) followed by Buliisa (12.1%), and Mayuge (4.9%) [12]. Notably, 7% of baseline participants with periportal fibrosis also exhibited chronic hepatitis-like or early cirrhosis-like disease [12]. The high prevalence of schistosomiasis-related morbidity remained despite the study area having at least over 10 rounds of school-based and community-based MDA with praziquantel preceding the cohort baseline [12].

The health system in Uganda is organized into four levels [23], starting with village health teams that support health promotion, basic service delivery, and community engagement. Health Centre IIs serve as the first formal point of care through outpatient and outreach services, while Health Centre IIIs provide broader preventive, promotive, and curative care. Community hospitals (Health Centre IVs) and general hospitals expand these services and perform more complex procedures such as surgery and blood transfusion. Regional referral hospitals deliver specialized clinical services, teaching, and research, and national referral hospitals provide the highest level of diagnostic and clinical care. Patients are expected to begin care at lower levels and move upward through referrals when needed.

### Study design and participant recruitment

A one-day workshop was held in each of the three SchistoTrack study districts, Pakwach, Buliisa, and Mayuge on the 21^st^, 22^nd^, and 25^th^ October 2024 respectively, to assess health care worker training needs for schistosomiasis morbidity management. The District Health Office (DHO) was asked to identify key health personnel involved with patient care when referred from peripheral clinics or outreaches, especially for severe cases of schistosomiasis. Participants from health facilities of varying levels within the Uganda health care system, were selected due to their likely involvement and having seen patients affected by schistosomiasis. Invited participants who were targeted included sonographers, medical doctors, clinical officers, laboratory personnel, and inpatient care nurses. Representation from across these levels ensured inclusion of diverse professional cadres and experiences relevant to schistosomiasis surveillance and clinical management. In addition to the participants, representatives from the central Ministry of Health, local district government management, and University of Oxford were present, and included, district health officers, local council 5 (LC5) politicians, a consultant gastroenterologist experienced with severe schistosomiasis morbidity management, the SchistoTrack project managers from Oxford and Uganda, and the principal investigators from Oxford and Uganda.

### Structure of the health worker workshops

The workshops were structured into six sessions that included in order: 1) presentations on the current burden of schistosomiasis in Uganda and an introduction to the SchistoTrack cohort, 2) a clinical case report by an expert clinician, 3) an interactive session on patient case studies from the SchistoTrack cohort, 4) mapping of patient pathways, 5) anonymous participation and feedback, and 6) a demonstration of schistosomiasis diagnosis.

Representatives from the Ministry of Health, and the SchistoTrack team presented updates on schistosome transmission, disease burden, and mass drug administration in Uganda as of 2024. This was followed by an introduction to the SchistoTrack cohort focusing on summarising the prevalence of infections and key conditions measured in the cohort, presenting information relevant only to the district where the workshop was held. The infections and conditions presented included *S. mansoni*, malaria, hepatitis B, Human immunodeficiency virus (HIV), and *S. mansoni*-related morbidity including gastrointestinal conditions (ever vomited blood), palpable (enlarged) spleen and or, liver, moderate anaemia, severe anaemia, and periportal fibrosis (See Supplementary File S1, S1 Table). The presentations were followed by questions from district participants and answers by the central Ministry of Health and SchistoTrack teams. The main aim of the presentations was to give background of schistosomiasis problems seen in each district.

After the presentations, a complex case definition of a hepatosplenic patient was presented by an experienced medical officer (JBO) from Pakwach, (the district with the highest burden of severe liver disease [14]), as an illustration of a case management and possible over-management of a patient (See Supplementary File S1, S1 Methods). This discussion and complex case study was not formally analysed but used to initiate discussions around patient scenarios before moving to an interactive session.

For interactive sessions, within small groups, participants were presented with patient cases from the SchistoTrack study, which were examined in January-February 2024 in the respective districts. These cases, spanning different age groups, illustrated the full spectrum of intestinal schistosomiasis morbidity, including both positive and negative findings, as well as associated comorbidities, lifestyle factors, laboratory results, imaging features, and clinical outcomes. Detailed examples of the case studies are provided in the Supplementary File S1 (Figures S1-S5).

Participants were then divided into groups of 5-7 people and moderated by central Ministry of Health staff or SchistoTrack team members, to map out the patient pathways for an assigned patient case study. Each group comprised a variety of expertise, including laboratory technicians, clinicians, and sonographers, who might not have interacted regularly within the existing health system structure. The group discussions were guided by pre-set facilitator questions (See Supplementary File S1, S2 Methods) which focused on current health system procedures for handling a patient from triage through diagnosis, treatment, and follow-up. The discussions also identified some training needs and resources needed in each district for effective management of intestinal schistosomiasis.

Participants then used flip charts to make group presentations of their patient pathways, identified training and resource needs, and proposed solutions for intestinal schistosomiasis management relevant to their local context. Discussions to revise each presentation regarding how the patient should be handled were guided by experts from the SchistoTrack team and the central Ministry of Health. After the patient pathway mapping exercise, facilitators observed that participants did not feel comfortable openly discussing their training and resource needs, perhaps due to fear of showing their inadequacy to their colleagues. Hence, participants were invited to write down their needs, questions and proposed solutions on pieces of paper and submit them anonymously. The questions were read to the wider group and discussed.

Finally, participants were taken through demonstrations that included Kato-Katz microscopy for identification of schistosome eggs and ultrasound-based assessment of schistosomiasis-related fibrosis performed by SchistoTrack laboratory technicians (BN) and sonographers (VA), respectively. In addition, praziquantel, the recommended treatment for schistosomiasis, was shown to participants some of whom had not seen the medicine. No feedback was recorded from these demonstrations.

## Data analysis

Workshop discussions were documented through notes taken in English. All transcripts and notes were read multiple times to develop familiarity with the collected data. The data were then manually coded using qualitative thematic analysis [24]. An inductive approach [25] was employed to group participant quotes into codes, allowing themes to emerge during the coding process rather than being predetermined [24]. Two authors (PMI and GFC) validated the codes through inter-coder agreement. Recurrent codes were consolidated into three overarching themes, which were reported as research findings. The full set of codes and themes is presented in Supplementary File S1, S2 Table.

## Results

### Study demographics

Workshop participant characteristics are presented in Table 1. A total of 105 participants attended the workshops, drawn from Pakwach (23.8%), Buliisa (36.2%), and Mayuge (40.0%) districts. There were more males than females, with males comprising 68.6% (72/105) of participants. Nursing Officers were the most common cadre, accounting for 36.1% (38/105) of participants, followed by laboratory staff (22.8%) and clinical officers (20.0%). Most participants were employed at Health Centre IIIs, representing 49.5% (52/105) of the participants, compared with fewer participants from Health Centre IIs and IVs.

**Table 1.** Study participants by district, level of health facility, and cadre (N = 105)

| Characteristic | Category | N (%) |
| --- | --- | --- |
| **District** | Pakwach | 25 (23.8) |
|  | Buliisa | 38 (36.2) |
|  | Mayuge | 42 (40.0) |
| **Gender** | Male | 72 (68.6) |
|  | Female | 33 (31.4) |
| **Age group (years)** | 25 - 34 | 37 (35.2) |
|  | 35 - 44 | 43 (41.0) |
|  | 45 - 54 | 20 (19.0) |
|  | 55+ | 5 (4.8) |
| **Job cadre** | Medical Officers | 15 (14.3) |
|  | Clinical Officers | 21 (20.0) |
|  | Nursing Officers | 38 (36.1) |
|  | Laboratory Staff (Technologists, Technicians, and Assistants) | 24 (22.8) |
|  | Public Health & Vector Control Officers | 5 (4.8) |
|  | Sonographers | 2 (1.9) |
|  | Ministry of Health and Administrative leadership roles (e.g. District Health officers, Local council 5) | 4 (3.8) |
|  | Other specialized staff (Researchers) | 2 (1.9) |
| **District employment\*** |  |  |
| **Pakwach** | Health Centre III | 11 (68.8) |
|  | Health Centre IV | 5 (31.2) |
| **Buliisa** | Health Centre II | 1 (4.0) |
|  | Health Centre III | 18 (72.0) |
|  | Health Centre IV | 6 (24.0) |
| **Mayuge** | Health Centre II | 2 (7.7) |
|  | Health Centre III | 23 (88.5) |
|  | Health Centre IV | 1 (3.8) |

District employment\* - Health facility levels where participants worked by district

### Case definitions of schistosomiasis

Across the three study districts, health workers demonstrated notable gaps in knowledge regarding schistosomiasis case definitions and disease classification. They struggled to distinguish between current infection and chronic morbidity. Liver complications were frequently misclassified as cirrhosis or hepatitis, reflecting a lack of standardized diagnostic approaches. One participant from Buliisa asked, *“On behalf of the clinicians, what is cirrhosis vs schistosomiasis?”* This confusion was compounded by the scarcity of trained sonographers with experience in schistosomiasis morbidity sonography and limited familiarity with the Niamey protocol - a WHO standardized, ultrasound-based guideline for assessing hepatosplenic schistosomiasis, and a lack of simplified, standardized reference tool for grading fibrosis from mild to severe.

Few health staff had experience with abdominal palpations. Discussions in Buliisa revealed that few participants knew how to perform abdominal palpations and interpret ultrasound examinations, while in Pakwach only one person reported having performed palpations. Interpretation of ultrasound findings varied widely, with participants noting a disconnect in terminology and understanding between sonographers, laboratory technicians, and clinicians. A participant from Buliisa observed that, *“There is a gap between clinicians, sonographers, and the laboratory in the language they use [to grade schistosomiasis-related morbidity].”* A senior clinician in Mayuge emphasized the importance of communication with sonographers: *“When you do an ultrasound request, meet the sonographer for more insight regarding the results. Ask the sonographer what you want to see, don’t just say do a normal or an abnormal scan.”*

The discussions also revealed that it was unclear among health workers about how to distinguish between severe and non-severe cases. For instance, a medical officer -from a private clinic-shared a case of a schistosomiasis patient who still had normal liver function but presented with advanced symptoms such as portal hypertension, ascites, and upper gastrointestinal bleeding. Because there is no clear system for grading varices, the team decided to manage the patient with surgery and regular endoscopic banding. This case showed the need for clearer guidance on how to identify and manage severe cases.

> *‘… There is no system for grading varices, so we opted for surgery.’ (Clinician, Mayuge)*

Diagnostic practices for current infection were limited. Laboratory technicians predominantly relied on the wet mount technique. As one participant from Buliisa explained, *“The current method for diagnosing schistosomiasis is a wet mount technique…the reagents are expensive, they [health facilities] don’t feel they get enough cases and are worried about expiry [of reagents], and there is a low suspicion index [of schistosomiasis] so we are not asked to look for it [schistosomiasis].”* Another participant from Mayuge confirmed, *“Laboratory people currently diagnose with a wet prep.”*

Table 2 presents key questions and proposed solutions raised through anonymous participation regarding case definitions, across the three study districts.

**Table 2:** Schistosomiasis case definitions.

| Theme | Key questions raised | Proposed solutions |
| --- | --- | --- |
| **Case definitions of schistosomiasis** | What clinical and diagnostic criteria define a schistosomiasis morbidity case? | 1). Develop simplified diagnostic protocol and classification guidelines for schistosomiasis morbidity.<br>2). More guidance is needed on how to interpret ultrasound findings to diagnose either severe or moderate schistosomiasis. |
|  | Should treatment be initiated based on ultrasound evidence in the absence of eggs? | Guidelines unavailable. Participants were advised that such patients can still be treated with praziquantel to address current infections. |
|  | How can clinicians differentiate schistosomiasis-related liver fibrosis from cirrhosis or hepatitis? | Develop and implement structured in-service training, mentorship and refresher courses for clinicians, laboratory staff, and sonographers. |
|  | What are the implications of using the wet mount technique rather than Kato-Katz for surveillance? | No solution provided |
|  | How can we get training on Kato-Katz technique and reagent to use in the laboratory? | Advocate for the inclusion of schistosomiasis and schistosomiasis-related morbidity in the Health Management Information System. |
|  | How can the lower-level facilities diagnose the cases of schistosomiasis in the history taking or physical examinations? | 1). Rural health facilities in endemic areas should be equipped with POC-CCA tests, so that schistosomiasis can be detected at Health Centre II level and below.<br>2). Introduce abdominal palpations at lower health facilities. |
|  | What is specifically needed during the assessment of the patient with schistosomiasis -clinical signs-before requesting a scan? | Develop simplified diagnostic protocol and classification guidelines for schistosomiasis morbidity. |
|  | What are the clinical signs and symptoms for schistosomiasis, especially in early stages?<br>How do you diagnose early and late of bilharzia? | 1). Develop and avail practical job aids in health facilities for use.<br>2). Conduct joint workshops with clinicians, sonographers, and other laboratory technicians to harmonize diagnostic language across cadres. |
|  | What treatment is given in late or severe case of bilharzia and in adults and children? | No solution provided |

### Patient pathways for *S. mansoni*-related morbidity management

During the interactive group discussions on mapping patient pathways, participants discussed current procedures within the health systems including how patients are managed from triage, to diagnosis, to treatment, to referral and to follow-up. It was evident that patient pathways for schistosomiasis morbidity care exist in most health facilities but are not standardized. For instance, one group in Mayuge district explained that: *‘At our triage point we take oxygen saturation and temperature… We take a detailed history: If they [patients] vomited blood, if they have abdominal pain, previous medical history of malaria, HIV…’*

The participants however noted that health workers had limited written protocols and standard referral and feedback procedures. It was also reported that many patients fail to reach referral centers due to high transport costs while others self-refer to private or faith-based hospitals.

> *‘How shall the facility get feedback on the ultrasound result when the client is referred to another facility?’ (Participant, Mayuge)*
>
> *‘Patients are often referred without feedback, and no one tracks if they reach the next facility.’ (Participant, Mayuge)*
>
> *‘Sometimes [patients] are given appointment days to go to see a doctor in Pakwach - but cannot go due to lack of transport.’ (Participant, Pakwach)*

Triage and clinical assessment were described as weak, with minimal physical examination such abdominal palpation. While private hospitals were reported to have the equipment and capacity to manage schistosomiasis-associated morbidity, they lacked standardized clinical guidelines and references to classify schistosomiasis-related morbidity. As a result, care often depends on the experience and judgment of the individual clinician.

> *‘The private hospital has a sonographer, they [doctors] don’t have a reference to determine if something is abnormal or not, they go on with what the sonographer says.’ (Participant, Mayuge)*

There is no mechanism for systematic morbidity monitoring, and schistosomiasis cases were not captured in the National Health Management Information System (HMIS).

> *‘The ministry of health HMIS does not capture schistosomiasis, it is lumped together under gastrointestinal disorders… so we cannot track the disease.’ (MoH, Pakwach)*

Additionally, it was highlighted that there was a need to agree on the categorization and risk management of emergency and chronic cases. A schistosomiasis expert attending the workshop in Mayuge advised, *‘We need to categorise risk in patients even without symptoms.’*

During the discussions, participants suggested using existing supplies and resources from other programs, such as blood and emergency kits from the maternity sections, to support schistosomiasis case management when emergencies arise. This idea reflects an opportunity for integrated service delivery in primary health facilities rather than having parallel, disease-specific systems, enhancing sustainability of schistosomiasis morbidity programs.

> ‘*All the maternity units have emergency kits…can we provide those emergency kits [to manage schistosomiasis emergency cases and replace later] …’ (Participant, Pakwach)*

Table 3 summarises key questions and proposed solutions arising from anonymous participation on patient pathways for managing schistosomiasis-related morbidity across the three study districts.

**Table 3:** Schistosomiasis patient pathways.

| Theme | Key questions raised | Proposed solutions |
| --- | --- | --- |
| **Patient pathways for *S. mansoni*-related morbidity management** | Who should conduct triage and initial assessment for suspected schistosomiasis cases? | Ensure triage is conducted by trained clinical personnel using structured forms. |
|  | What protocols should guide referral and follow-up for severe morbidity cases? | Introduce referral feedback forms and integrate them into HMIS reporting. |
|  | Should presumptive treatment be provided when diagnostics are unavailable? | Participants were advised that such patients can still be treated with praziquantel to address current infections. |
|  | How can facilities ensure continuity of care after referral? | Establish follow-up registers to track patient outcomes post-referral. |
|  | What provisions are needed for emergency management of severe morbidity? | Provide emergency kits and ensure consistent supply of blood products and IV fluids. |

### Capacity building and resources needed for effective management of schistosomiasis

Health workers highlighted significant gaps, including limited skills in interpreting Kato-Katz and ultrasound results, the absence of training curricula for lower-level staff, insufficient mentorship, and the lack of integrated continuing education programs on schistosomiasis morbidity.

> *‘We need comprehensive training on complications of schistosomiasis, both emergency and chronic. (Sonographer, Buliisa workshop)*
>
> *‘Orientation on the Kato-Katz technique and provision of reagents are necessary.’ (Participant, Mayuge)*
>
> *‘Sonographers need uniform training in grading fibrosis and communicating findings to clinicians.’ (Participant, Buliisa)*
>
> *‘Health workers should be empowered through continuous mentorship and continuing medical education.’ (Ministry of Health representative, Pakwach)*

Shortages or the lack of praziquantel within health facilities was reported. Clinicians expressed uncertainty over dosage regimens, retreatment intervals, and management of severe cases, outside of an MDA setting. There was uncertainty about whether praziquantel should be administered to patients already presenting with advanced schistosomiasis morbidity, such as bleeding varices or periportal fibrosis.

> *‘…Should we use praziquantel in case of serious complication but no current infection?’ (Participant, Mayuge)*

During discussions in Mayuge, facilitators clarified that while praziquantel effectively kills adult flukes, there is still need to confirm if it can reverse established fibrosis or portal hypertension. Moreover, in cases of active gastrointestinal bleeding, its use has been suspected to carry additional risks but this still needs further investigations. In Buliisa, one participant asked*, “What if the ultrasound scan shows fibrosis but the laboratory results are negative for eggs, should I still treat?”* Facilitators explained that no formal guidelines currently exist for such cases. They noted that laboratory tests, particularly the wet mount method-which is mainly used in the study areas, is a weak method to detect schistosome eggs. In these situations, diagnosis is primarily based on ultrasound findings, which may indicate past exposure. Participants were advised that such patients can still be treated with praziquantel to address current infections. Participants called for updated treatment guidelines, including praziquantel in the national essential and medical stores lists, and clear protocols for case management.

> *‘Would you clearly guide us on how and when to use praziquantel?’ (Participant, Buliisa)*
>
> *‘We are working to put PZQ on the essential medicine list.’ (MoH representative, Buliisa)*

Participants emphasized the need for strengthening laboratory capacity, and structured, practical training in diagnostic and clinical management techniques, alongside mentorship and inclusion of schistosomiasis morbidity in professional development programs at all levels of care.

Table 4 summarises key questions and proposed solutions arising from anonymous participation on resources needed for effective management of schistosomiasis across the three study districts.

**Table 4:** Capacity building and resources needed for effective management of schistosomiasis.

| Theme | Key questions raised | Proposed solutions |
| --- | --- | --- |
| **Capacity building and resources needed for effective management of schistosomiasis** | What specific training content is needed for schistosomiasis morbidity management? | Develop short module courses on schistosomiasis diagnosis, management, and ultrasound interpretation.<br>Train sonographers and clinicians on a schistosomiasis morbidity protocol (Niamey protocol) |
|  | How can training be institutionalized within existing continuing medical education structures? | Integrate schistosomiasis morbidity modules into national in-service training curricula for clinicians, sonographers, and laboratory technicians. |
|  | What mentorship models are effective for rural facilities? | Establish mentorship networks linking regional specialists with peripheral health facilities |
|  | Why is praziquantel not routinely available in health facilities? | Include praziquantel in the national essentials medicine and medical stores procurement list for all levels of care. |
|  | What is the correct PZQ dosing protocol in cases of chronic morbidity? | No guidelines in this area. |
|  | How can facilities access diagnostic reagents and equipment? | Provide laboratories with Kato-Katz reagents, functional microscopes, and ensure each district has at least one ultrasound unit. |

## Discussion

Recent WHO policy emphasises the urgent need to improve schistosomiasis morbidity control and strengthen health facility-based case management beyond traditional infection-focused strategies [1]. We conducted an assessment to understand health worker needs and capacity for schistosomiasis morbidity management in three high-burden rural districts of Uganda. Through interactive workshops that combined presentations, clinical case reviews, patient-pathway mapping, ultrasound demonstrations, and anonymous feedback, we engaged clinicians, laboratory personnel, nurses, sonographers, and senior health management teams drawn from across all levels of the district health system. Our findings revealed substantial gaps in case definitions, patient management pathways as well as a lack of specialized personnel (sonographers and radiographers) and access to essential resources such as praziquantel, ultrasound equipment, and standardized morbidity assessment tools. Mass drug administration of praziquantel in endemic countries has been successful in reducing infection burden over the past 30 years [26]. In our study districts, participants reported that access to praziquantel remains inadequate, as praziquantel is rarely stocked in public health facilities. Clinicians were unsure of dosage regimens, diagnostic tools to rely on, and management of severe cases outside an MDA setting. This is in line with similar studies conducted in schistosomiasis-endemic regions of SSA that have reported that the integration of schistosomiasis control measures into primary health care systems is often constrained by limited praziquantel availability, and insufficient training among primary health care staff [27–29].

Praziquantel is listed on the WHO essential medicine list [30], and as reported by the schistosomiasis national program representative (HK) during our workshops, Uganda is working towards adding it on the Essential National Drug List and incorporating it into the medicine quantification exercises that allocate medicines to facilities, representing an important step toward strengthening schistosomiasis case management.

### Need for standardised case definitions of schistosomiasis morbidity

Growing recognition of the substantial morbidity associated with schistosomiasis has led the WHO to recommend passive case detection by routine health services, and that health facilities ensure access to praziquantel for all affected individuals, irrespective of age [1]. To translate these recommendations into effective practice, there is a need for better case definitions. Historically, infection intensity has been used as a proxy for schistosomiasis morbidity. In most resource-limited settings, including our study districts, the direct wet smear technique is the primary method used to detect current schistosomiasis infection. Although this technique is rapid and requires no reagents, it has considerably lower sensitivity compared with other diagnostic methods [28, 31, 32]. More sensitive approaches such as the Kato-Katz technique, the Point-of-Care Circulating Cathodic Antigen (POC-CCA) test, and ultrasound are often unavailable due to the additional time required to perform these tests [33], cost, prioritization of other diseases such as malaria and human immunodeficiency virus (HIV), shortages of reagents [31], and limited technical expertise [34]. King and colleagues [35], among others, [11, 36] have emphasized that defining schistosomiasis solely by current infection is inadequate, as disease manifestations can persist long after parasites have been cleared. According to WHO guidance on routine health information systems and facility data for neglected tropical disease [37], a suspected case of intestinal schistosomiasis in endemic areas is defined as a person presenting with non-specific abdominal symptoms, blood in stool, or hepatosplenomegaly, while a confirmed case requires the microscopic detection of schistosome eggs in stool. The International Classification of Diseases [ICD]-11, adopted in 2019 and effective from 2022, provides a global standard for recording and comparing mortality and morbidity data, and classifies schistosomiasis into acute and chronic forms [37]. Despite these references, operational criteria remain vague. For instance, hepatosplenic schistosomiasis presents considerable heterogeneity, ranging from mild symptoms to severe, life-threatening disease [38]. Its progression is often subtle, with few clinical signs until the first episode of upper gastrointestinal bleeding occurs, while advanced stages can lead to liver failure [39]. Also, recent evidence [11, 12] suggests that morbidity is disconnected from current infection so that WHO suggestions of using current infection to confirm actual cases of schistosomiasis is flawed. The current acute-chronic categorization is overly simplistic and should be revised to encompass the full spectrum of disease with, as we have learned from our workshops, criteria needed to manage patients where clinicians need information on what is a mild, moderate, or severe case rather than only a group of suspected symptoms generally related to schistosomiasis.

At present, laboratory and presumptive diagnosis, as well as case management, largely depend on the individual expertise of and ultimately exposure to a high number of cases for health personnel [27, 28]. In this study, health workers expressed the need for clearer guidance on clinical and diagnostic criteria for intestinal schistosomiasis morbidity, particularly regarding how to respond when ultrasound reveals fibrosis in the absence of eggs, how to differentiate schistosomiasis-related liver disease from conditions such as hepatitis or cirrhosis, and how to appropriately identify early versus advanced disease at lower-level health facilities. Participants also noted a lack of consensus when interpreting ultrasound results between sonographers, laboratory technicians, and clinicians, including which expertise to prioritize and how best to translate ultrasound findings for clinicians unfamiliar with reading images but responsible for managing patients. Participants recommended the development of an in-service training module that covers schistosomiasis diagnosis, clinical management, and ultrasound interpretation using the Niamey protocol, along with instruction in abdominal palpation for use in lower-level health facilities. They further suggested that these trainings be conducted jointly with clinicians, sonographers, and laboratory technicians to promote harmonized diagnostic language across different professional cadres.

Although disease burden and the number of severe cases encountered varied across our study districts, variation in burden did not necessarily translate into variation in needs. For instance, for severe cases, the challenge was not a lack of knowledge per se; health workers recognised these presentations because they resembled other emergency cases requiring stabilisation encountered in routine health services. Instead, the challenge was lack of operational guidelines with a shortage of clinical resources and non-inclusion of schistosomiasis in HMIS reports.

Schistosomiasis control initiatives in SSA have largely relied on vertical approaches [31, 40]. While such programmes can deliver rapid disease control, they often struggle with sustainability due to poor integration into the broader health system [28, 31, 41–44]. There remain needs for improved country ownership of schistosomiasis morbidity and inclusion of schistosomiasis praziquantel on the National Essential Dug list so that funds can be allocated to purchase Praziquantel from national medical stores, as well as clearer codes for documenting severe schistosomiasis cases within HMIS especially in the absence of current infection. Less severe cases posed greater uncertainty, as their definitions are more ambiguous and require clearer guidance. Using data previously collected and analysed in the SchistoTrack Cohort [14], it was possible to develop clear, straight forward definitions of severe schistosomiasis especially in cases where patients present with factors such as hypersplenism, upper gastrointestinal bleeding, ascites, and or severe anaemia. This definition, complemented with targeted health education for less severe presentations, may be a starting point for a standardised case definition within primary health systems.

### Need for clear patient pathways

This study revealed fragmented patient pathways, unstandardised triage, minimal physical and laboratory examination, and unreliable referral systems; patterns widely observed in other schistosomiasis-endemic countries [40, 45, 46]. The clinical management of hepatosplenic schistosomiasis is inherently complex, and mainly focuses on minimising variceal bleeding, and reducing mortality [47]. The management of the disease is further complicated in the presence of coinfection with viral hepatitis and cirrhosis, and the different presentation of pathology between children/adolescents and adults [47]. Despite these challenges, systematic evaluations comparing different treatment strategies and pathways are unavailable, and standardised clinical management guidelines have not been established [47]. Given the variability of hepatosplenic schistosomiasis, as shown in our patient case studies, a uniform management strategy is unlikely to be suitable for all clinical presentations of hepatosplenic schistosomiasis [47].

Case management and patient pathways work with passive case detection, which is usually triggered by patients taking action to seek care based on a number of factors [40]. Factors such as the healthcare setting, the level of clinical expertise available, and social and clinical circumstances play an important role in determining the most appropriate management strategy [47]. Previous studies have shown that referral to other facilities brings more burden of time and costs for the patients [40]. This finding was supported in our study, which reported that many patients fail to reach referral centers due to high transport. Additionally, there are only few experts and public health workers trained to identify cases of severe intestinal schistosomiasis especially in SSA [34]. In our workshops, we had only two trained sonographers drawn from a private hospital who were familiar with diagnosing hepatosplenic disease; the rest of the participants lacked familiarity with the WHO Niamey ultrasound protocol [48] for assessing schistosomiasis-related morbidity, and even had limited experience with abdominal palpations. This limited expertise, concentrated in a private facility, further restricts access to care, as affordability becomes a barrier for patients in need of diagnostic services. It is therefore important to consider the ability or willingness of a patient to adhere to long-term follow-up, and available health care infrastructure and resources when designing a management plan. A recent review by Frischer et al. [46], demonstrated that care-seeking and referral pathways for neglected tropical diseases, including severe intestinal schistosomiasis morbidities [49] remain incomplete. Guidelines local to each endemic country are needed that outline clear referral pathways and integration with other health service platforms. The management pathways need to be simple, with the extent of standardisation depending on the complexity of the case and whether chronic or emergency care is needed.

### Need for practical approaches to strengthen health system capacity

To achieve schistosomiasis elimination targets, control programs must be complemented by broader, integrated approaches that embed schistosomiasis control within the primary health care systems [50, 51]. In our study, health workers proposed using supplies from other programs, such as blood and emergency kits from maternal health services, to support schistosomiasis case management during emergencies. They also highlighted coordination with better-equipped stakeholders, including private clinics, as a practical solution for managing schistosomiasis-related morbidity by building capacity for public facilities that currently have inadequate skilled personnel in this area and lack most of the essential tools.

To help forecast needs, track progress, and assess the impact of interventions, the WHO has now issued guidance on how schistosomiasis data should be captured and reported [20]. Member States are expected to submit national data through standardized channels: the Epidemiological Reporting Form and Joint Reporting Form for preventive chemotherapy outcomes. In addition, passive surveillance data from health facilities, such as the number of patients seen, should be incorporated into monthly reports to national health information systems [20]. WHO also recommends a set of minimum core indicators for schistosomiasis data collection and reporting, which support monitoring, evaluation, programme management, and country progress tracking toward the 2021–2030 road map targets [20]. In Uganda, the Ministry of Health has revised the National Essential Health Care Package [23] to promote universal coverage through a multisectoral and collaborative approach. The updated package includes services for managing schistosomiasis in public health facilities, alongside broader health system strengthening measures, particularly improvements in referral networks and emergency transport. Our findings can help to further promote the prioritisation of the health system indicators in light of considering which indicators are most practical for clinical decision making.

## Limitations

Some participants were hesitant to openly share their training and resource needs, which may have limited the depth of our discussions though we had hoped to mitigate this limitation through anonymous feedback. Furthermore, the workshops did not include all types of health workers in Uganda, which may limit the generalizability of the findings, particularly regarding knowledge of schistosomiasis morbidity and its management. Nonetheless, we consider the perspectives obtained to be broadly representative. Given that we held large workshops, audio recording was not possible so we may have missed some comments in the manual transcriptions.

## Conclusion

This study has shown that standardised schistosomiasis diagnostic and grading reference tools, together with adequate and regular training of health workers, are essential for effective recognition and management of the disease. These measures are necessary if health facilities are to provide routine treatment and management of schistosomiasis, as recommended by WHO. We anticipate that the framework for engaging health workers here and the patient case studies provided can be used in other countries to conduct needs-based assessments to improve schistosomiasis-morbidity management. Insights gathered in this study also provide a foundation for context-appropriate clinical guidance and training programmes.

## Acknowledgements

We are grateful to our study participants, and the SchistoTrack teams, especially the nurses, sonographers, and laboratory technicians. We also like to thank the Uganda Ministry of Health, and the local district leaders. Special thanks also to the Oxford team for everyday discussions, and feedback.

## Data availability

All data in the present work are contained in the manuscript and supplementary files.

## Ethics approvals

This study was approved by Oxford Tropical Research Ethics Committee (OxTREC 509-21), Vector Control Division Research Ethics Committee of the Uganda Ministry of Health (VCDREC146), and Uganda National Council for Science and Technology (UNCST HS 1664ES).

## Author contributions

Conceptualization: GFC. Data curation: GFC, PMI, BN, LW, JBO, NU, MO, JN, HK, PB, VA, CKO, NBK. Formal analysis: PMI. Investigation: PMI, LW, BN. Methodology, visualization: PMI, NBK, GFC. Writing – original draft: PMI, GFC. Validation: PMI, GFC. Writing – review and editing: GFC, PMI, BN, LW, JBO, NU, MO, JN, HK, PB, VA, CKO, NBK. Funding acquisition and supervision: GFC. Resources: GFC.

## Supporting information

Supplementary File S1: S1 Methods: An example of hepatosplenic case management by an expert clinician. S2 Methods: Pre-set facilitator questions used during the workshops to guide discussions on mapping outpatient pathways. S1 Table: Infections and conditions measured within the SchistoTrack Cohort by District in 2024. S2 Table: The full set of codes and themes generated from workshop discussions. Figure S1: Case study P1. Figure S2: Case study P2. Figure S3: Case study B3. Figure S4: Case study M1. Figure S5: Case Study M3.

