## Supplementary File for "Government health care worker training needs for schistosomiasis morbidity management"

**Supplementary File S1 for:**

### Government health care worker training needs for schistosomiasis morbidity management

Phyllis Munyiva Isaiah[1], Betty Nabatte[2], Lauren Wilburn[1], John Bosco Oryema[3], Noah Ukumu[3], Morris Okumu[4], Juma Nabhonge[5], Hilda Kyarisiima[2], Prudence Beinamaryo[2], Victor Anguajibi[6], Christopher K. Opio[7], Narcis B. Kabatereine[2], Goylette F. Chami[1]*

*

#### Supplementary Methods

**S1 Methods**: An example of a hepatosplenic case management by an expert clinician

The patient had experienced progressive abdominal distension over a two-year period, which had worsened in the preceding three days, prior to observation. On general examination, findings were documented and monitored over several days to assess the evolving clinical picture. Laboratory results showed neutropenia, with additional tests- liver function (Aspartate Aminotransferase (AST)/ alanine transaminase (ALT) tests), as well as renal function parameters performed to evaluate possible organ involvement. The management plan included emergency blood transfusion, administration of furosemide, tranexamic acid, propranolol, and spironolactone, with insertion of a urethral catheter to monitor urine output and assess renal function. The clinician emphasized the role of a plant-based protein diet to reduce hepatic metabolic load while maintaining adequate nutrition. A plasma expander was used to address hypovolemia. Notably, the patient passed melena stools predominantly at night, a finding discussed in relation to portal hypertension and variceal bleeding risk. The clinician also highlighted that the AST-to-Platelet Ratio Index (APRI) score can be used to assess liver fibrosis severity. Ultimately, a review of the referral system revealed that the patient had been lost to follow-up.

**S2 Methods**: Pre-set facilitator questions used during the workshops to guide discussions on mapping out patient pathways

1. What are the first steps for patient triage?
2. What would be the provisional diagnoses?
3. How do you make this decision? (name any tests/examinations you would you do?)
4. Who makes this decision? Who works with who?
5. What would be the differential diagnoses?
6. How do you make this decision? (name any tests/examinations you would you do?)
7. Who makes this decision? Who works with who?
8. What are some key issues that persons quality of life/ what are the patients concerns?
9. What can you address with resources available?
10. What can you not address with resources available?
11. Should the patient be referred? If so what would be the reason?
12. What are some solutions if you cannot refer the patient?
13. How do you communicate the findings with the patients?
14. Do you come across any issues here?
15. Would you follow up with the patients?
16. How would you contact them?
17. How long after would you follow them up?
18. Are there any issues with patient follow up?
19. Is there anything you are uncertain of?

#### Supplementary Tables

**S1 Table**: Infections and conditions measured within the SchistoTrack Cohort by District in 2024

| District | Condition | Overall Prevalence (%) | Adults (>18 Years) (%) | Children (<18 Years) (%) |
|---|---|---|---|---|
| **Pakwach** | *S. mansoni i*nfection | 53.0 (766/1445) | 38.5 (295/767) | 69.5 (471/678) |
| | ***S. mansoni* -related morbidity** | | | |
| | Gastrointestinal condition (vomiting blood ever) | 1.4 (20/1415) | 2.7 (20/750) | 0 |
| | Palpable (enlarged) spleen | 11.8 (167/1414) | 12.4 (93/750) | 11.1 (74/664) |
| | Palpable (enlarged) liver | 4.5 (63/1414) | 4.8 (36/750) | 4.1 (27/664) |
| | Moderate anaemia (8-10.9 g/dL Hb) | 36.6 (527/1439) | 27.2 (208/766) | 47.4 (319/673) |
| | Severe anaemia (< 8 g/dL Hb) | 8.3 (120/1439) | 6 (46/766) | 11 (74/673) |
| | Periportal fibrosis | 26.4 (376/1425) | 38.1 (288/755) | 13.1 (88/670) |
| | Malaria | 25.8 (354/1374) | 12.3 (89/726) | 40.9 (265/648) |
| | Hepatitis B | 5.9 (85/1439) | 9.8 (75/766) | 1.5 (10/673) |
| | Human immunodeficiency virus (HIV) | 4.6 (65/1402) | 8 (60/749) | 0.8 (5/653) |
| **Buliisa** | *S. mansoni i*nfection | 46.6 (514/1102) | 33.0 (190/576) | 61.6 (324/526) |
| | ***S. mansoni* -related morbidity** | | | |
| | Gastrointestinal condition (vomiting blood ever) | 0.6 (7/1079) | 1.2 (7/566) | 0 |
| | Palpable (enlarged) spleen | 7.1 (76/1070) | 4.7 (26/555) | 9.7 (50/515) |
| | Palpable (enlarged) liver | 1.3 (14/1070) | 1.6 (9/555) | 1.0 (5/515) |
| | Moderate anaemia (8-10.9 g/dL Hb) | 21.6 (238/1103) | 12.5 (72/578) | 31.6 (166/525) |
| | Severe anaemia (< 8 g/dL Hb) | 2.3 (25/1103) | 1.2 (7/578) | 3.4 (18/525) |
| | Periportal fibrosis | 14.2 (156/1096) | 22.6 (130/574) | 5 (26/522) |
| | Malaria | 23.3 (258/1108) | 10.2 (59/580) | 37.7 (199/528) |
| | Hepatitis B | 6 (66/1103) | 9.7 (56/578) | 1.9 (10/525) |
| | Human immunodeficiency virus (HIV) | 5.2 (56/1084) | 8.4 (48/570) | 1.6 (8/514) |
| **Mayuge** | *S. mansoni i*nfection | 22.1 (174/789) | 14.8 (64/433) | 30.9 (110/356) |
| | ***S. mansoni* -related morbidity** | | | |

| | | | | |
|---|---|---|---|---|
| | Gastrointestinal condition (vomiting blood ever) | 0.3 (2/760) | 0.5 (2/421) | 0 |
| | Palpable (enlarged) spleen | 3.2 (24/759) | 2.4 (10/414) | 4.1 (14/345) |
| | Palpable (enlarged) liver | 1.1 (8/759) | 1.2 (5/414) | 0.9 (3/345) |
| | Moderate anaemia (8-10.9 g/dL Hb) | 12.1 (95/785) | 8.6 (37/431) | 16.4 (58/354) |
| | Severe anaemia (< 8 g/dL Hb) | 0.6 (5/785) | 0.9 (4/431) | 0.3 (1/354) |
| | Periportal fibrosis | 9.8 (76/772) | 14.4 (61/425) | 4.3 (15/347) |
| | Malaria | 14.2 (111/784) | 5.1 (22/430) | 25.1(89/354) |
| | Hepatitis B | 2.9 (23/784) | 4.9 (21/431) | 0.6 (2/353) |
| | Human immunodeficiency virus (HIV) | 3.8 (30/782) | 6.3 (27/430) | 0.9 (3/352) |

**S2 Table**: The full set of codes and themes generated from workshop discussions

| Theme | Codes |
|---|---|
| **Case definitions of schistosomiasis** | i. Inconsistent criteria for active vs. chronic, and severe vs. non-severe schistosomiasis<br>ii. Wet mount technique preferred due to lack of Kato-Katz reagents<br>iii. Confusion between schistosomiasis-related liver disease, cirrhosis, and hepatitis<br>iv. Limited knowledge of Niamey Protocol<br>v. Few clinicians perform abdominal palpations<br>vi. No standard reference for abnormal ultrasound findings |
| **Patient pathways for *S. mansoni*-related morbidity management** | i. Triage is often handled by untrained staff<br>ii. Absence of standardized triage and referral forms<br>iii. Weak feedback mechanisms between referring and receiving facilities<br>iv. Limited follow-up for chronic schistosomiasis cases<br>v. Inadequate emergency supplies and blood transfusion<br>vi. Referral non-adherence due to high transport costs<br>vii. Informal reliance on private clinics for ultrasound services<br>viii. HMIS does not capture schistosomiasis as a distinct category |
| **Capacity building and resources needed for effective management of schistosomiasis** | i. Limited diagnostic and clinical skills in Kato-Katz and ultrasound interpretation<br>ii. Absence of training curricula for lower-level health workers on diagnosis and management of schistosomiasis and its related morbidity<br>iii. Lack of mentorship and on-site refresher training<br>iv. Need for integrated continuing education programs on schistosomiasis morbidity<br>v. Unreliable access to praziquantel in public health facilities<br>vi. Absence of Kato-Katz reagents and functional microscopes<br>vii. Lack of ultrasound machines in public health facilities<br>viii. Limited expertise in schistosomiasis management |

#### Supplementary Figures

Below are examples of patient case studies presented during the workshops across the three study districts (Figure S1- Figure S5). These cases were examined in January-February 2024 within the SchistoTrack Study.

| **Figure S1:** Case study P1 | |
|---|---|
| **Demographics** | 10-19-year-old male student. |
| **Medical History** | Self-reported history of malaria, typhoid, gut worms in the past three years. |
| **Other history** | Non-smoker, non-drinker |
| **Clinical symptoms** | Reported symptoms of fever, loss of appetite, cough, flu and bloody diarrhoea in the past month. Visited a government health care facility in the past month and took antimalarials and non-praziquantel deworming medicines. |
| **Blood and stool analyses (Diagnostics)** | - Malaria RDT trace positive, positive for *P. falciparum* 144 /µL<br>- 312 *S. mansoni* eggs per gram of stool (mild intensity). Negative for other soil-transmitted helminths<br>- Normal stool (Bristol stool type 4), faecal occult blood trace positive<br>- Haemoglobin 13.3 g/dL, WBCs 7 x$10^9$/L<br>- Negative for Hepatitis B and HIV |
| **Ultrasonography and palpations** | - Palpations: liver and spleen were not palpable. No other clinical symptoms were noted.<br>- Ultrasound scan: Liver patterns C. Liver had a sharp edge. No other abnormalities were noted |
| **Referral** | None |
| **Findings** | NA |

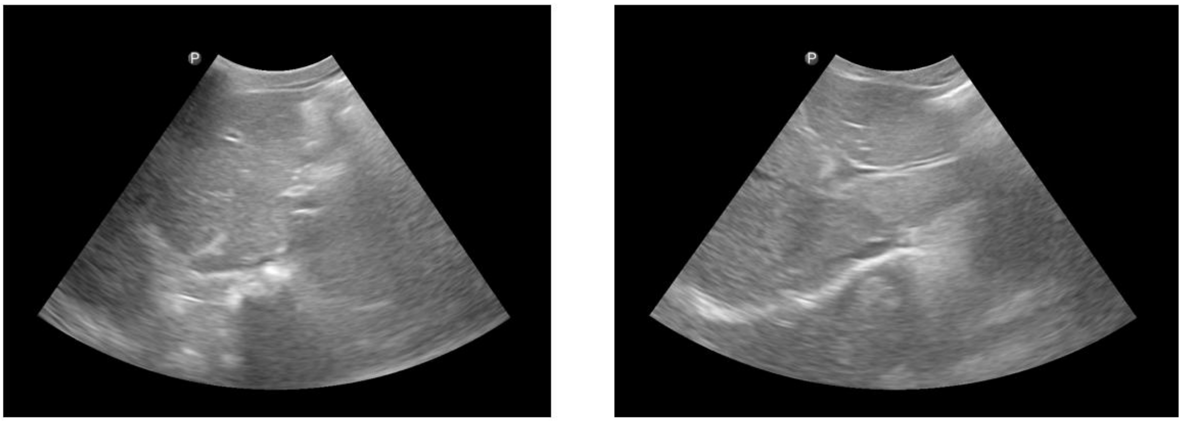

C1 C2

**Patient Case P1**: ***showing periportal fibrosis liver patterns C***
**Pattern C**: rings with no echogenic centre or pipe stems (long stretched vessels)**.**

| **Figure S2:** Case study P2 | |
|---|---|
| **Demographics** | 60-69-year-old female with no current or previous occupation. |
| **Medical History** | Previously reported taking praziquantel and herbalism.<br>History of vomiting blood as early as 2002.<br>Received a blood transfusion but has not vomited blood in the past year. |
| **Other history** | Non-smoker, non-drinker, not pregnant. |
| **Clinical symptoms** | Reported recent symptoms of headache, fever, cough. In the past month reported a mass in the abdomen, collapse/fainting, and severe abdominal pain. Has taken amoxicillin and omeprazole in the past month. |
| **Blood and stool analyses (Diagnostics)** | - Negative for malaria, soil-transmitted helminths, and *S. mansoni*<br>- Normal stool (Bristol type 4), no blood visualised in stool, faecal occult blood negative<br>- Haemoglobin 9.2 g/dL, WBCs 3.4 x 109 / L<br>- Negative for HIV and hepatitis B |
| **Ultrasonography and palpations** | - Palpations: Liver was not palpable; spleen was palpable with a firm consistency. No other clinical signs were recorded<br>- Ultrasound scan: Liver patterns D and E, liver had a sharp edge, splenic varies and gall stones observed |
| **Referral** | None |
| **Findings** | NA |

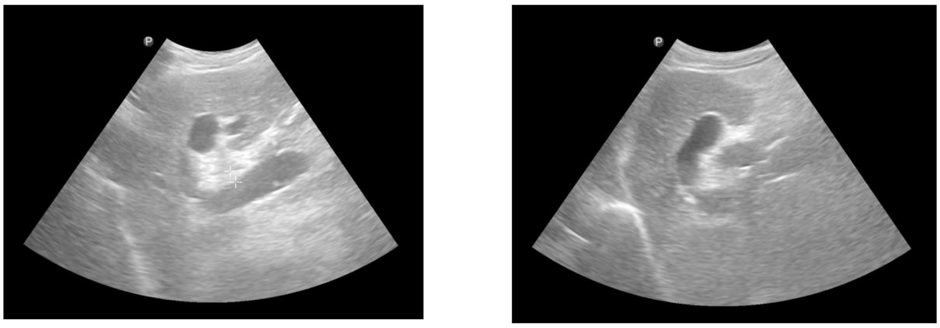

D E

***Patient Case P2: showing liver patterns D and E***

***Pattern D:*** Thickness around the main portal vein

**Pattern E:** Blocked vessel that should have no lumen.

| **Figure S3:** Case study B3 | |
|---|---|
| **Demographics** | 30-39-year-old male fisherman. |
| **Medical History** | Self-reported history of pertussis, eczema and typhoid. |
| **Other history** | Smokes hand-rolled cigarettes every day and drinks large sachets of homebrewed alcohol every day. |
| **Clinical symptoms** | Reported symptoms in the past month of severe abdominal pain and described a mass in the abdomen. Went to seek care at a private health clinic and took antimalarials and paracetamol |
| **Blood and stool analyses (Diagnostics)** | - Negative for malaria, soil-transmitted helminths and *S. mansoni*<br>- Normal stool (Bristol stool type 4), no visible blood in stool, faecal occult blood negative.<br>- Haemoglobin 13.8 g/dL, WBCs 1.4 x $10^9$/L<br>- HIV negative<br>- Positive for Hepatitis B for the past two years, did not self-report a history of previous positive hepatitis test. |
| **Ultrasonography and palpations** | - Palpations: liver and spleen not palpable, no other clinical symptoms<br>- Ultrasound scan: Liver patterns B and C, liver had a sharp edge, no other abnormalities. |
| **Referral** | None |
| **Findings** | NA |

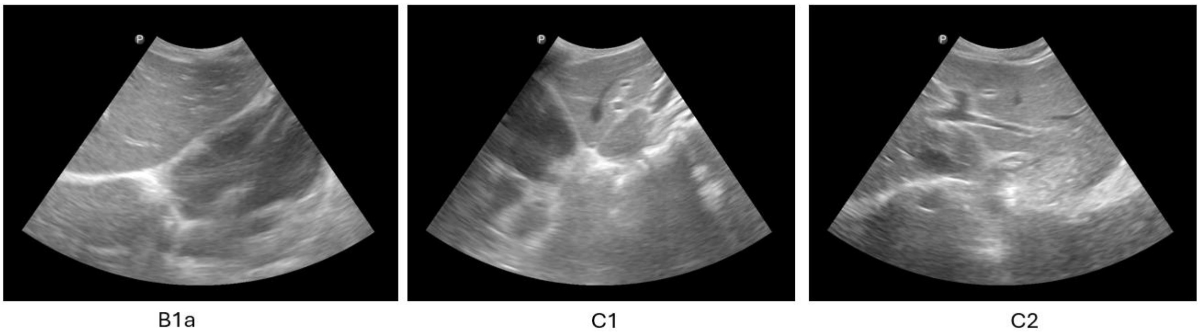

***Patient case B3: showing liver patterns B and C***

**Pattern B**: small dots (starry sky) or large, solid white dots (flying saucers)

**Pattern C**: rings with no echogenic centre or pipe stems (long stretched vessels)**.**

| **Figure S4:** Case study M1 | |
|---|---|
| **Demographics** | 30-39-year-old female with no occupation. |
| **Medical History** | No self-reported history of infections or conditions.<br>Reports of a previous history of upper gastrointestinal tract bleeding, the first occurrence happened in 2021. In the last year, one instance of vomiting blood, three months prior to observation. The patient did not receive a blood transfusion for this. |
| **Other history** | Non-drinker, non-smoker. Not pregnant. |
| **Clinical symptoms** | The patient had headaches, epistaxis (nosebleeds) and a mass in the abdomen in the past month. |
| **Blood and stool analyses (Diagnostics)** | - Negative for malaria, soil-transmitted helminths, and *S. mansoni.*<br>- Faecal occult blood negative, no blood visualised in stool<br>- Slight diarrhoea (Bristol stool type 5)<br>- Haemoglobin 9.6 g/dL, WBCs in normal range 4.7 x $10^9$/L<br>- Negative for HIV and Hepatitis B |
| **Ultrasonography and palpations** | - Palpations: firm liver, hard spleen and abdominal swelling<br>- Ultrasound scan: Normal liver (Pattern A), gall bladder stones, normal liver surface, splenic varices, moderate splenic infarction<br>- Other endomyocardial fibrosis |
| **Referral** | Referred to the health centre 4 for further management. |
| **Findings** | Splenomegaly and portal hypertension. |

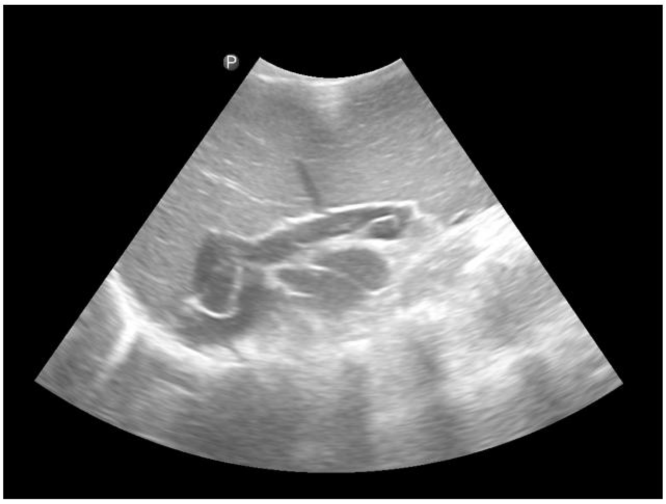

A

***Patient Case M1: showing a normal liver (Pattern A)***

**Pattern A**: no signs of periportal thickening.

| **Figure S5:** Case Study M3 | |
|---|---|
| **Demographics** | 50-59-year-old male sustenance farmer. |
| **Medical History** | No self-reported infections or conditions. |
| **Other history** | Non-drinker, non-smoker. |
| **Clinical symptoms** | No symptoms reported in the last year. |
| **Blood and stool analyses (Diagnostics)** | - Negative for malaria, soil-transmitted infections and *S. mansoni*<br>- Normal stool (Bristol stool type 4) no visible blood in stool<br>- Faecal occult blood positive<br>- Haemoglobin 11 g/dL, WBCs 1.3 x $10^9$/L<br>- Negative for Hepatitis B and HIV |
| **Ultrasonography and palpations** | - Palpations: liver not palpable, spleen palpable and a hard texture<br>- Ultrasound: Liver patterns C and E, Liver had a sharp edge.<br>- Gastro-oesophageal varices were reported |
| **Referral** | None |
| **Findings** | NA |

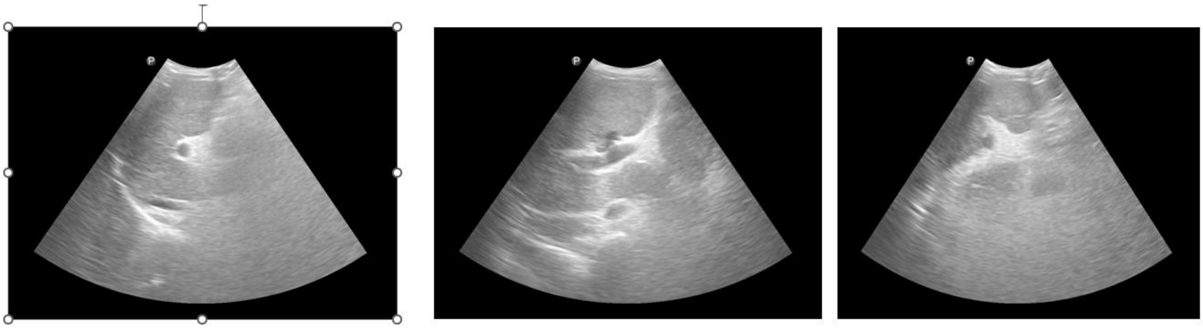

C1 C2 E

***Patient Case M3: showing liver patterns C and E***

**Pattern C**: rings with no echogenic centre or pipe stems (long stretched vessels)

**Pattern E:** Blocked vessel that should have no lumen.
